# Anemia and iron metabolism in COVID-19: A systematic review and meta-analysis

**DOI:** 10.1101/2020.06.04.20122267

**Authors:** Petek Eylul Taneri, Sergio Alejandro Gómez-Ochoa, Erand Llanaj, Peter Francis Raguindin, Lyda Z. Rojas, Beatrice Minder Wyssmann, Doris Kopp-Heim, Wolf E. Hautz, Michele F. Eisenga, Oscar H. Franco, Marija Glisic, Taulant Muka

**Affiliations:** Bahcesehir University, Faculty of Medicine Public Health Department Istanbul/Turkey; Public Health and Epidemiological Studies Group, Cardiovascular Foundation of Colombia, Floridablanca, Colombia; Public Health Research Institute, University of Debrecen, Hungary; Doctoral School of Health Sciences, Faculty of Public Health, University of Debrecen, Hungary; Institute of Social and Preventive Medicine (ISPM), University of Bern, Bern, Switzerland; Swiss Paraplegic Research, Nottwil, Switzerland; Research Group and Development of Nursing Knowledge (GIDCEN-FCV), Research Institute, Cardiovascular Foundation of Colombia, Floriadablanca, Santander, Colombia; Public Health & Primary Care Library, University Library of Bern, University of Bern, Bern, Switzerland; Department of Emergency Medicine, Inselspital, Bern University Hospital, University of Bern, Bern, Switzerland; Department of Internal Medicine, Division of Nephrology, University Medical Center Groningen, University of Groningen, Groningen, The Netherlands

**Keywords:** anemia, hemoglobin, ferritin, iron, covid-19, prognosis

## Abstract

Iron metabolism and anemia may play an important role in multiple organ dysfunction syndrome in Coronavirus disease 2019 (COVID-19). If confirmed, this has important implications for the more than 1.62 billion people estimated to have anemia globally. We conducted a systematic review and meta-analysis to evaluate biomarkers of anemia and iron metabolism (hemoglobin, ferritin, transferrin, soluble transferrin receptor, hepcidin, haptoglobin, unsaturated iron-binding capacity, erythropoietin, free erythrocyte protoporphyrine, and prevalence of anemia) in patients diagnosed with COVID-19, and explore their prognostic value. Six bibliographic databases were searched up to May 5^th^ 2020. We included 56 unique studies, with data from 14,044 COVID-19 patients (59 years median age). Pooled mean hemoglobin and ferritin levels in COVID-19 patients across all ages were 130.41 g/L (95% Confidence Interval (CI), 128.42; 132.39) and 673.91 ng/mL (95% CI, 420.98; 926.84), respectively. Hemoglobin levels decreased with advancing age and increasing percentage of comorbid and critically ill patients, while levels of ferritin increased with increasing male proportion and mean hemoglobin levels. Compared to moderate cases, severe cases had lower pooled mean hemoglobin [weighted mean difference (WMD), –4.21 (95% CI –6.63; –1.78)] and higher ferritin [WMD, –730.55 ng/mL (95% CI 413.24; 1047.85)]. A significant difference in mean ferritin level of 1027.23 ng/mL (95% CI 819.53; 1234.94) was found between survivors and non-survivors, but not in hemoglobin levels. No studies provided information on anemia or other biomarkers of interest. Future studies should explore the impact of iron metabolism and anemia and in the pathophysiology, prognosis, and treatment of COVID-19.

## INTRODUCTION

Infection with severe acute respiratory syndrome coronavirus 2 (SARS-COV-2) often results in Coronavirus disease 2019 (COVID-19), a disease that endangers disproportionately the elderly, those with pre-existing chronic conditions such as cardiovascular disease, diabetes mellitus and hypertension(1, 2). If deteriorating, Covid-19 can lead to sepsis, septic shock, and multiple organ dysfunction syndrome, with mechanical ventilation or extracorporeal membrane oxygenation having low therapeutic efficacy(3). The pathophysiological background underlying deterioration and low efficacy of common treatments is unclear.

Most patients with COVID-19 who require intensive care will develop an atypical form of the acute distress respiratory syndrome (ARDS) with preserved lung gas volume (4), suggesting hypoxia due to physiological processes other than alveolar dysfunction may play a role in the prognosis of the disease(5). Disturbed iron metabolism may be one such affected process. Indeed, recent data show that COVID-19 patients tend to present decreased hemoglobin levels indicating the presence of anemia, and pathologically increased levels of ferritin. A study of 67 COVID-19 patients in Singapore reported that during their course in an intensive care unit (ICU), patients developed more profound and significantly lower hemoglobin levels, compared to patients not admitted to ICU (6). Another study in elderly patients hospitalized for COVID-19 found that most patients had hemoglobin levels lower than the normal range, but did not find significant differences in hemoglobin levels between survivors and non-survivors. However, follow-up was incomplete for half of the patients(7). In a report of 5700 patients hospitalized for COVID-19 in the New York City area, ferritin levels were pathologically high, a finding in line with previous studies from China (8, 9). Both anemia and hyperferritenemia, regardless of the underlying pathology, are strong predictors of mortality(10, 11). Anemia could be the result of iron-restricted erythropoiesis arising from alterations in iron metabolism. Increased ferritin levels could be indicative of a strong inflammatory reaction in COVID-19 or related to viral entry into the human body and its impact on iron metabolism(12, 13). Iron is an essential micronutrient for both humans and pathogens(14). The innate immune response could restrict iron availability during infections to deprive the pathogen of it, a mechanism that would also lead to anemia (15, 16). Anemia, in turn, reduces oxygen delivery to the tissue and may thus play an important role in the development of multi organ failure. Therefore, it is crucial to understand the relation between anemia, iron metabolism and progression of COVID-19, and whether these associations differ by age, sex and presence of chronic conditions.

We thus conducted a systematic review and meta-analysis of available observational evidence to (i) quantify the mean levels of hemoglobin, ferritin and other biomarkers of iron metabolism in COVID-19 patients, (ii) explore whether the levels would differ by age, sex presence of chronic conditions and severity of COVID-19, and (iii) whether these biomarkers could have clinical and/or prognostic utility in COVID-19.

## METHODS

We conducted a systematic review and meta-analysis according to a recent published guideline and reported according to PRISMA (Preferred Reporting Items for Systematic Reviews and Meta-Analyses) guidelines17). The protocol was registered in PROSPERO and is available in https://www.crd.york.ac.uk/prospero/display_record.php?RecordID=180670.

### Data source and strategy

We searched MEDLINE (National Library of Medicine, US), EMBASE (Elsevier, Netherlands), Web of Science (Clarivate Analytics, US), Cochrane (Cochrane Collaboration, UK), the WHO COVID-19 database and Google Scholar (Google, Inc., US) to identify relevant articles. We used search terms related to COVID-19 infection and SARS-CoV-2 virus, and several markers of anemia and iron storage and metabolism, including hemoglobin, ferritin, transferrin, soluble transferrin receptor, hepcidin, haptoglobin, unsaturated iron-binding capacity, erythropoietin and free erythrocyte protoporphyrin. We also included search terms related to clinical progression of COVID-19 such as prognosis, severity, ICU admission, mortality, risk factors, clinical features, clinical characteristics or predictors. We searched the databases from inception until May 05^th^ 2020. We limited our search to human studies and restricted our analysis to articles published in English. The detailed search strategies are presented in **eAppendix 1**.

### Study selection and eligibility criteria

All observational studies (e.g., cross-sectional, cohort, and case-control studies), except for case reports and case-series, were included. We included studies that reported on the levels of the biomarkers of iron metabolism, erythropoietin or hemoglobin levels, the prevalence of anemia in COVID-19 patients, or their levels by the clinical outcome of patients with COVID-19. Outcomes of interest include disease severity, admission to intensive care unit, mechanical ventilation, and mortality across all age groups. Studies examining the association between the biomarkers of interest and risk of COVID-19 complications (e.g., admission to intensive care unit, death) were also included.

### Data extraction

Two independent reviewers screened the titles and abstracts according to the selection criteria. We recorded data on the author’s name, study location, study design, sample size, clinical characteristics, laboratory results, disease severity, and outcome in a data extraction form. The form was developed, piloted, and discussed within the review group before full data extraction. All laboratory values were extracted as reported and then converted to conventional units based on the US National Institute of Standards and Technology conversion factors. For studies reporting only median and ranges (interquartile range, range, and maximum-minimum values), we converted these values into mean and standard deviation (Hozo, 2015).

### Risk of Bias assessment

The quality of included studies was assessed by two authors independently using the Newcastle-Ottawa Scale for case-control, cross-sectional and cohort studies, as applicable. A third author adjudicated in case consensus was not reached. The scale was developed for non-randomized and observational studies and assesses quality in three broad categories, namely, selection of study groups/participants, comparability of the study groups/participants, and the assessment of exposure/outcome of interest. Quality was assessed on a 10-point scale and classified as good quality (9–10 points), fair quality (6–8), and poor quality (< 6).

### Data synthesis and analysis

Based on the extracted data of each study, we computed pooled means and standard deviations for each biomarker and anemia prevalence for all COVID-19 patients, and weighted mean difference in levels of biomarkers between severe vs. moderate cases, and survivors vs. non- survivors. For the adult population, anemia was defined according to World Health Organization (WHO) guidelines (hemoglobin levels for males < 130 g/L and for females < 120 g/L). We pooled mean difference by using a random-effects model (DerSImonian and Laird, 1986). Geographical location, age group (pediatric vs. adult), the mean age of the study population, gender distribution, percentage of comorbidities and proportion of patients admitted to ICU were pre-specified as characteristics for assessment of heterogeneity and were evaluated using stratified analyses and univariate random effects meta-regression. Publication bias was appraised using funnel plot as well as Egger’s test for assessing asymmetry. We used STATA 15.1 (Statacorp, Texas, US, 2017) for all the analyses. We only calculated 2-tailed tests. A p-value < 0.05 was considered significant.

## RESULTS

### Study Identification and Selection

1302 unique citations were identified of which 178 were selected for full-text evaluation. Of those, 56 observational studies comprising 14,044 individuals diagnosed with COVID-19 were included for further analysis (**Figure 1**). References of the 56 included studies can be found in the Supplemental Material. Forty-nine studies provided information on blood hemoglobin, and eighteen studies provided information on serum ferritin. We did not find studies reporting on other markers of iron metabolism or providing information on anemia according to WHO age and sex-specific criteria or on erythropoietin. Detailed characteristics of the included studies can be found in **Supplemental Table S1**. In brief, besides three studies conducted in the USA (n = 2) and Italy (n = 1), all identified studies were conducted in Asia. The majority of studies were conducted in an adult population (n = 39, 69.6%), fifteen studies (24.7%) included both pediatric and adult patients, and two studies included pediatric patients only. The median age of the mean population age was 59 years (IQR 48–63.7 years), the median proportion of males was 60.3% (IQR 49.5–67.5%) and the median percentage of patients having comorbidities was 46% (IQR 32.8–49).

**Figure 1.**
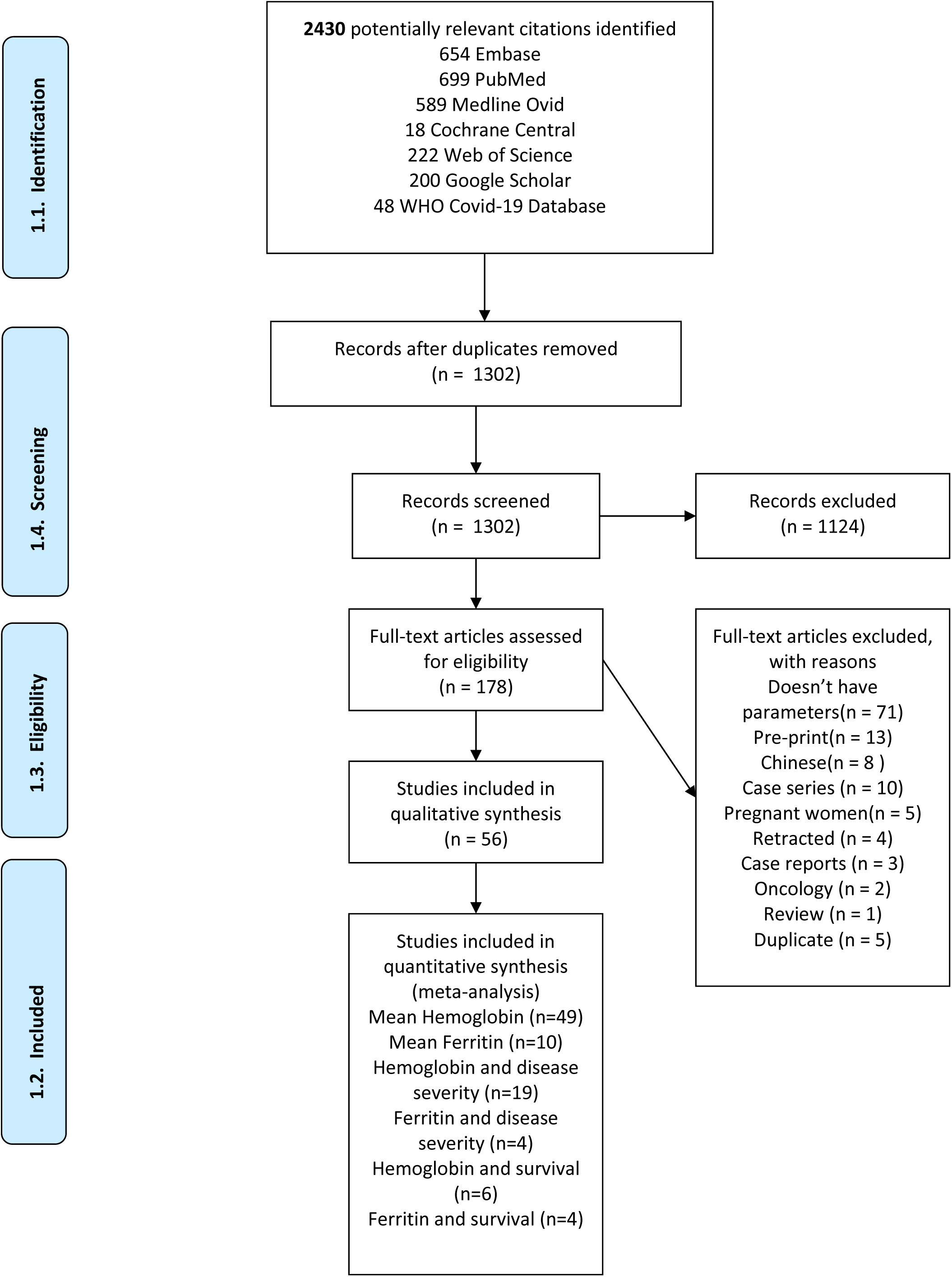
Flowchart of included studies

### Serum Hemoglobin Levels

Based on findings from 49 observational studies, including 8,828 individuals, pooled mean hemoglobin level was 130.41 g/L (95% Confidence Interval (CI) 128.42; 132.39; *I^2^* = 98.3%, *P*-value for heterogeneity< 0.001), **Table 1 and eFigure 1**. Compared to studies including individuals with mean age below 50.33 (overall median age of all studies), studies with mean population age above 50.33 years had higher mean hemoglobin levels [133.033 g/L (95% CI 131.20; 135.145) vs. 127.15 g/L (95% CI 125.13; 129.17)] (**Supplementary Table S2**). Similarly, after regressing the mean age per study and mean serum hemoglobin levels, a significant linear trend was observed, with mean serum hemoglobin levels decreased with increasing age (**Figure 2a**). Also, studies including exclusively adult populations, had lower mean serum hemoglobin levels in comparison to studies including both adults and pediatric individuals [129.15g/L (95% CI 126.58; 131.71) vs. 133.59g/L (95% CI 131.04; 136.414)] (**Supplemental Table S2**). The stratified analysis showed lower mean hemoglobin levels in studies including patients with a higher prevalence of comorbidities, especially of type 2 diabetes, and with a higher proportion of patients admitted to ICU, while no differences were observed by the proportion of males and the percentage of survivors by the end of the study (**Supplementary Table S2**). Fitting regression lines showed similar results, albeit no significant for type 2 diabetes (**Supplementary Table S2, Figure 2b and eFigure 5**). Huang et al. reported reduction in hemoglobin levels in 38.2% of patients hospitalized for COVID-19, but did not specify the definition of decreased hemoglobin(18); while Wang et al reported decreased hemoglobin level (< 110g/L) in 19.23% of the study population admitted to hospital(1). In contrast, Xiu et al studied asymptomatic patients and reported none of the cases had decreased hemoglobin levels (19), albeit not defining the cut-off of decreased levels.

**Table 1.** Characteristics of studies included in meta-analysis of mean hemoglobin and ferritin levels and the main results. Bubble plots with fitted meta-regression line of (a) the mean serum hemoglobin levels and mean age among 49 observational studies; (b) the mean serum hemoglobin levels and mean percentage of patients hospitalized at intensive care unit among 37 observational studies; the mean serum ferritin levels and mean age and percentage of male population among 9 observational studies respectively and (c/d) and mean ferritin levels regressed against mean hemoglobin levels based on findings from 7 observational studies (e). Circles are sized according to the precision of each estimate with larger bubbles for more precise estimates.

| OUTCOME | Eligible studies | Participants |  |  | Location |  |  |  |  | Results of meta-analysis |  |  |  |
| --- | --- | --- | --- | --- | --- | --- | --- | --- | --- | --- | --- | --- | --- |
|  | Unique studies, no. | Total | Median (IQR), no. | Age, median (IQR), years | Europe | North America | South America | Asia-Pacific | Middle East | Overall mean (95% CI) | P value | I2 for heterogeneity | P value for heterogeneity |
| **Hemoglobin, g/L** | 49 | 8,828 | 85 (40.5-192) | 50.33 (45.1-58.2) | 1 | 1 | --- | 47 | --- | 130.41(128.42;132.39) | <0.001 | 98.3% | <0.001 |
| **Ferritin, ng/mL** | 10 | 5,565 | 1971 (65.5-219.25) | 56.35 (49.7-58.5) | --- | 1 | --- | 9 | --- | 673.91 (420.98;926.85) | <0.001 | 99.7% | <0.001 |
| **Ferritin<sup>1</sup>, ng/mL** | 9 | 5,555 | 174 (91-237.5) | 56.7 (53.4-58.5) | --- | 1 | --- | 8 | --- | 737.58 (539.93;935.24) | <0.001 | 99.2% | <0.001 |
| <sup>1</sup>excluding a single study focusing on pediatric population (Tan et al) |  |  |  |  |  |  |  |  |  |  |  |  |  |

**Figure 2.**
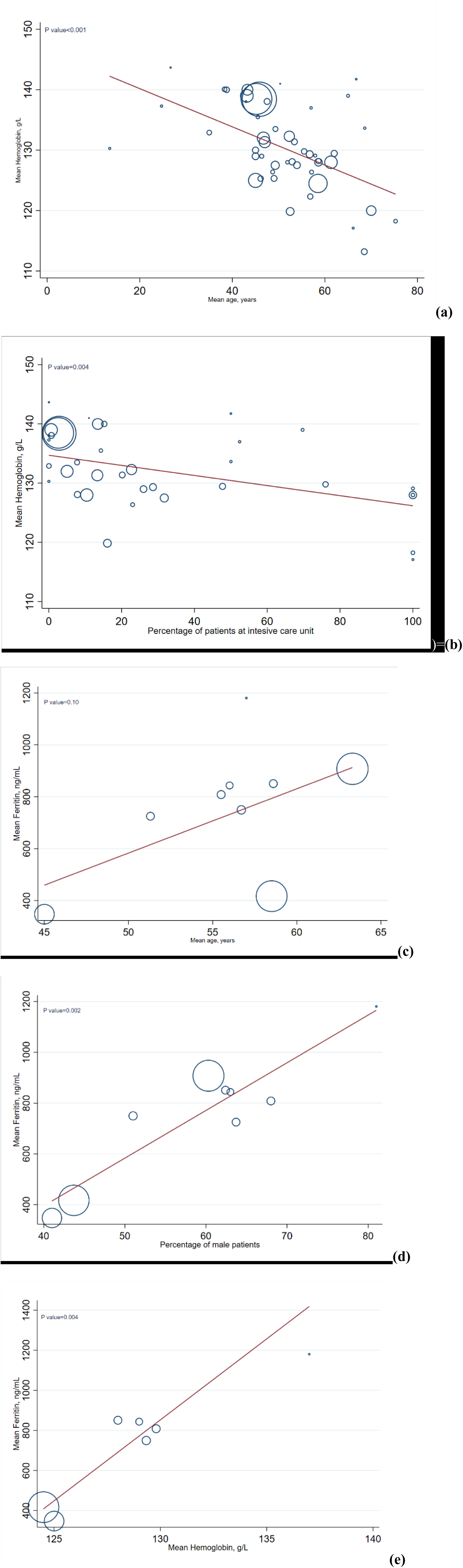
Meta-regression results

When comparing mean hemoglobin levels in severe and moderate cases (using data from 19 studies and 3,251individuals with COVID-19), severe cases had lower hemoglobin levels [weighted mean difference (WMD), –4.21 (95% CI –6.63; –1.78); *I^2^* = 62.6%, *P*-value for heterogeneity< 0.001] (**Table 2, eFigure 2**). When comparing pooled estimates from studies above (18.95%) vs. below median (≤18.95%) the percentage of patients at ICU, WMD was larger in studies above the median [WMD –8.03 (95% CI –9.95; 6.12) vs. –2.62 (95% CI –5.41; 0.16) respectively; p-value = 0.04] (**Supplemental Table S3**). Nevertheless, age, percentage of male population and percentage of comorbidities were not major sources of heterogeneity (**Supplemental Table S3, eFigure 6**). Based on retrospective data from 245 individuals with COVID-19, Liu et al. found that the unadjusted association between baseline hemoglobin levels and all-cause mortality during hospitalization was non-significant, and the odds ratio of death with increasing serum hemoglobin level was 0.98 (95% CI 0.96, 1.00, p = 0.05)(20). Similarly, based on data from six studies we did not find a significant difference in hemoglobin levels between survivors and non- survivors [WMD, 0.24 (95% CI –2.39; 2.88); *I^2^* = 0%, *P-*value for heterogeneity = 0.85] (**Table 1 and eFigure 2**).

**Table 2.** Meta-analysis of differences in mean hemoglobin and ferritin levels based on disease severity and vital status.

| Biomarker | Eligible studies | Participants |  |  |  |  | Results of meta-analysis |  |  |  |
| --- | --- | --- | --- | --- | --- | --- | --- | --- | --- | --- |
|  | Unique studies, no. | Total | No. of **deceased individuals** | Mean biomarker level and 95% CI in deceased individuals | No. of **survived individuals** | Mean biomarker level and SD in survived individuals | WMD (95%CI) | P value | I2 for heterogeneity | P value for heterogeneity |
| **Hemoglobin, g/L** | 6 | 1,012 | 315 | 127.06 g/L (124.89;129.22) | 697 | 125.77 g/l (121.64; 129.91) | 0.24 (-2.39;2.88) | 0.86 | 0% | 0.85 |
| **Ferritin, ng/mL** | 4 | 561 | 245 | 1587.68 ng/mL (1395.75; 1779.61) | 316 | 546.05 ng/mL (494.08; 598.01) | 1027.23 (819.53; 1234.94) | <0.001 | 38.3% | 0.18 |
| Biomarker | Unique studies, no. | Total | No. of severe Covid-19 cases | Mean biomarker level and 95% CI in severe individuals | No. of Moderate Covid-19 cases | Mean biomarker level and SD in moderate cases | WMD (95%CI) | P value | I2 for heterogeneity | P value for heterogeneity |
| **Hemoglobin, g/L** | 19 | 3,251 | 1,213 | 129.62 g/L ( 126; 133.24) | 2,038 | 133.76 g/L (130.51;137) | -4.21 (-6.63;-1.78) | <0.001 | 62.6% | <0.001 |
| **Ferritin, ng/mL** | 4 | 154 | 53 | 1409.34 ng/mL (990.9; 827.78) | 101 | 625.1 ng/mL (336.72; 913.47) | 730.55 (413.24; 1047.85) | <0.001 | 63.6% | 0.04 |

### Serum Ferritin Levels

Based on findings from ten observational studies, including 5,565 COVID-19 patients, pooled mean ferritin level was 673 ng/mL (95% CI 420.98; 926.85) (**Table 1 and eFigure 4**). When excluding a single study performed in a pediatric population, the mean pooled ferritin level was 737.58 ng/mL (95% CI 539.93; 935.24) (**Table 1** and **eFigure 4**). Due to the limited number of studies we were only able to perform subgroup analyses comparing pooled ferritin levels in studies below and above median age (56.7 years) and the proportion of the males (62.4%) and found no significant differences (**Supplementary Table S3**). Yet, when regressing the mean age per study and mean serum ferritin levels we observed a significant linear trend with mean serum ferritin levels increasing with advancing age and the proportion of the male population (**Figure 2c&d**); After removing the single study performed in a pediatric population the trend between mean ferritin levels and age became non-significant (**eFigure 7**). Furthermore, when fitting regression lines using data from 7 studies (in adult patients), we observed a positive significant linear trend between mean ferritin and hemoglobin levels (**Figure 2e**).

When pooling estimates from four studies and 154 individuals, the mean difference in serum ferritin was higher in severe COVID-19 individuals compared to moderate cases; [WMD, 730.55 (95% CI 413.24; 1047.85) (**Table 2 and eFigure 3b**). Finally, pooling the estimates from four observational studies and 561 individuals we found a large statistically significant difference in mean ferritin levels of 1027.23 ng/mL between non-survivors and survivors [ WMD, 1027.23 (95% CI 819.53; 1234.94) (**Table 1 and eFigure 3**). In line with this finding, Zhou et al., found in a univariable analysis that odds of in hospital death were higher among patients with ferritin levels above 300ng/mL compared to those with serum ferritin ≤399ng/mL(odds ratio was 9.10 (95% CI 2.04; 40.58, p = 0.0038). Indeed, levels of serum ferritin were elevated in non-survivors compared with survivors (562 ng/mL±492 ng/mL) throughout the clinical course, and were increased with disease deterioration(3). Similarly, in a study by Li et al., ferritin was significantly higher in severe cases (21).

### Quality of studies and Publication Bias

Six (10,7%) of the included studies were evaluated at low risk of bias, while the rest at medium risk of bias (Supplemental table S1 and S4). Due to the limited number of studies included in comparative analyses of severity and survival of COVID-19, we could only test for publication bias in studies focusing on hemoglobin levels and comparing severe vs. non-severe cases, and survivors vs. non-survivors and found no evidence for publication bias (**eFigure 7**).

## DISCUSSION

To the best of our knowledge, the current study is the first updated and comprehensive systematic review and meta-analyses acknowledging the potential clinical utility of anemia and iron metabolism in COVID-19. Based on data from 56 studies and 14,044 COVID-19 patients across all ages, we found a pooled mean hemoglobin level of 130.41 g/L, which decreased with older age, higher proportion of comorbid illness and severity. We also found pathological values of ferritin in most patients, a finding more prominent in men and elderly. Major differences in ferritin levels were reported between different levels of severity, and among patients who survived and those who did not. We did not find any study in COVID-19 patients investigating the levels of biomarkers of iron status other than ferritin, and prevalence of anemia according to age and sex- WHO cut-off.

The mean level of hemoglobin in COVID-19 patient across all-ages found in this review represents a borderline value regarding the value for anemia diagnosis in men as defined by WHO (< 130 g/L), with anemic mean levels observed in the elderly, and in severe cases. Indeed, a study in 339 hospitalized elderly COVID-19 patients found that the majority of patients had mean values of hemoglobin lower than normal(7). However, most of the studies did not report hemoglobin levels by sex, and none of the studies reported the prevalence of anemia based on age and sex-specific WHO cut-offs. Thus, it was challenging to provide an accurate picture of the burden of anemia in COVID-19 patients. The SASR-COV-2 infection is associated with high mortality, especially among the elderly over the age of 65+ and comorbid patients.

Hemoglobin concentration is one of the most important determinants of the oxygen-carrying capacity of the blood. Low hemoglobin in COVID-19 patients, especially on populations at risk of complications and mortality, could indicate that the patients could suffer from a decreased capability of hemoglobin to support the increased peripheral tissue demands for oxygen due to the hyper-metabolic states during infection. Indeed, significant complications of COVID-19 patients are sepsis, septic shock, and multiple organ dysfunction syndrome, with mechanical ventilation or extracorporeal membrane oxygenation having low efficacy in mitigating its impact and progression(3). According to recent evidence, COVID-19 patients do experience an atypical form of the acute distress respiratory syndrome with preserved lung gas volume(4), suggesting that the prognosis could depend on the ability of the human body to meet the oxygen demands of the peripheral tissues, which, if not met, may lead to hypoxia and ischemia. Studies have reported that anemia is associated with 2.6 times increased risk of mortality in chronic obstructive pulmonary diseases; the overall 90-day mortality among these patients with acute respiratory failure treated with invasive mechanical ventilation was 57.1% versus 25% in non-anemic patients(22, 23). A previous meta-analysis has shown that among a mixed population, independent of sex, age and cardiovascular diseases, anemia is associated with a 41% and 33% increased risk of all-cause mortality and cardiovascular mortality, respectively (11). A similar risk of mortality was also shown in two other meta-analyses comparing heart failure patients with and without anemia, or stroke patients with and without anemia(24, 25). Also, another meta-analysis has shown that lower baseline hemoglobin values in heart failure patients are associated with increased crude mortality rates (r = –0.396, p = 0.025)(24). In this meta-analysis we show that the severity of disease and prognosis of patients with COVID-19 might depend on lower hemoglobin levels as severe cases had significantly lower hemoglobin levels than moderate cases. Future prospective studies with complete follow-up are needed to confirm the impact of anemia in COVID-19 outcomes, and whether the incidence of low hemoglobin levels does predict mortality in these patients.

A striking finding of this meta-analysis is the pathological value of ferritin among COVID-19 patients, with significant differences between severe and moderate cases, and survivors and non-survivors. Ferritin is known to be elevated in inflammatory conditions, with hyperferritinemia being a key acute-phase reaction used by clinicians as a marker for therapeutic response (12). However, recent studies suggest that increased levels of circulating ferritin levels may not only reflect an acute-phase response, but also play a critical role in inflammation by contributing to the development of a cytokine storm(12). According to Shoenfeld et al., the clinical picture of the critical cases of COVID-19 resembles macrophage activating syndrome, which is commonly associated with high levels of ferritin or even a cytokine storm(26). H-chain of the ferritin could be important in activating macrophages to increase the secretion of inflammatory cytokines observed in COVID-19 patients(26). Another explanation for the increased levels of ferritin could be the role of iron metabolism in supporting the innate immune system to fight invading microorganisms. The innate immune system orchestrates control over iron metabolism as a response to viral infections. For viral replication, enhanced cellular metabolism and optimal iron levels within host cells are necessary(13, 27). Therefore, the innate immune system will react by decreasing the bioavailability of iron to limit the replication of the virus during the acute phase of infection. In these conditions, through interleukin-6 and Toll-like-receptor-4 dependent pathways, the levels of the liver-derived iron hormone hepcidin-the master regulator of iron homoestasis- could increase and block, the activity of the transporter ferroportin which carries iron out of the cells, and therefore decrease the amount of iron absorbed from the diet, causing cellular sequestration of iron (i.e., principally in hepatocytes, enterocytes, and macrophages)(27). Increased intracellular iron sequestration will lead to an upregulation of cytosolic ferritin which sequesters and stores iron to prevent iron-mediated free radical damage (27). The increased retention and storage of iron within ferritin in macrophages contribute to the characteristic fall in serum iron concentrations and an increase in serum ferritin concentrations as observed in an acute phase response(28). The net result will be a diminished iron availability for erythropoiesis and as a result further aggravation of anemia. Regardless of the etiology, serum ferritin is highly elevated in patients with COVID-19, and it seems warranted to assess whether serum ferritin could be used as a screening biomarker for the severity of the inflammatory state that patients with COVID-19 have.

Our systematic review highlights important gaps in the role of iron biomarkers other than ferritin in the prognosis of COVID-19. Of the included iron status parameters, only information on ferritin levels was available. Future studies should measure also other iron status parameters to establish whether iron-restricted erythropoiesis is inflicting anemia, and whether anemia is contributing to poor outcomes. Assessing the other stated parameters and interpreting them with respect to iron status in the setting of inflammation will be a challenging task. Therefore, the inclusion of soluble transferrin receptor (sTfR) could be of additive value for iron-restricted erythropoiesis, as elevated levels of sTfR reflect both erythroid activity and functional iron deficiency. The markers is known to be less affected by concomitant chronic disease or inflammation. Future prospective population-based and clinical studies are necessary to investigate the utility of using levels of hemoglobin levels and iron biomarkers for risk stratification and to identify patients who could benefit from early prevention strategies. Provided that anemia and altered iron metabolism play a role in COVID-19, public health strategies may be designed to protect this the population that could be at risk of COVID-19 complications.

This study has several limitations. First, considering we restricted our review to articles in English, we cannot rule out publication bias, which could limit our overall findings. Second, the interpretation of the findings should be based on the quality of the included studies. Despite most studies being of high quality, the data provided are mainly of cross-sectional nature. Third, there was heterogeneity in the definition of moderate and severe cases of COVID-19 patients and in the definition of comorbid patients, which could have contributed to the observed heterogeneity in our meta-analysis.

This meta-analysis suggests that hemoglobin and ferritin levels vary according to the severity of COVID-19 as well as age, gender and presence of comorbidity among COVID-19 patients. Whether hemoglobin and ferritin can be used for prognostic purposes, or have further implications for identifying novel treatment targets, needs further investigation

## Data Availability

Upon request to authors, individual data from the studies included in the review can be freely available.

## Acknowledgement

This paper is a contribution of the authors to the UN Decade of Action on Nutrition (2016–2025) https://www.un.org/nutrition/

## Conflicts of interest/Competing interests (include appropriate disclosures)

Authors declare no conflicts of interest

## Authors’ contributions

Study concept and design: MG and TM

Acquisition, collection, analysis, or interpretation of data: PET, SAGO, EL, PFR, LZR, BMW, DKH, WEF, MFE, OHF, MG and TM,

### Drafting of the manuscript

PET, SAGO, EL, PFR, LZR, OHF, MG and TM

### Critical revision of the manuscript

PFR, LZR, BMW, DKH and WEF

All authors gave final approval and agree to be accountable for all aspects of work ensuring integrity and accuracy.

## References

1. Wang L, Duan Y, Zhang W, Liang J, Xu J, Zhang Y, et al. Epidemiologic and Clinical Characteristics of 26 Cases of COVID-19 Arising from Patient-to-Patient Transmission in Liaocheng, China. Clin Epidemiol. 2020;12:12–387.

2. Yang J, Zheng Y, Gou X, Pu K, Chen Z, Guo Q, et al. Prevalence of comorbidities and its effects in coronavirus disease 2019 patients: A systematic review and meta-analysis. Int J Infect Dis. 2020;94:94–91.

3. Zhou F, Yu T, Du R, Fan G, Liu Y, Liu Z, et al. Clinical course and risk factors for mortality of adult inpatients with COVID-19 in Wuhan, China: a retrospective cohort study. Lancet. 2020;395(10229):1054–62.

4. Gattinoni L, Coppola S, Cressoni M, Busana M, Rossi S, Chiumello D. Covid-19 Does Not Lead to a “Typical” Acute Respiratory Distress Syndrome. Am J Respir Crit Care Med. 2020.

5. Lang M, Som A, Mendoza DP, Flores EJ, Reid N, Carey D, et al. Hypoxaemia related to COVID-19: vascular and perfusion abnormalities on dual-energy CT. Lancet Infect Dis. 2020.

6. Fan BE, Chong VCL, Chan SSW, Lim GH, Lim KGE, Tan GB, et al. Hematologic parameters in patients with COVID-19 infection. Am J Hematol. 2020.

7. Wang L, He W, Yu X, Hu D, Bao M, Liu H, et al. Coronavirus disease 2019 in elderly patients: Characteristics and prognostic factors based on 4-week follow-up. J Infect. 2020.

8. Richardson S, Hirsch JS, Narasimhan M, Crawford JM, McGinn T, Davidson KW, et al. Presenting Characteristics, Comorbidities, and Outcomes Among 5700 Patients Hospitalized With COVID-19 in the New York City Area. JAMA. 2020.

9. Chen T, Wu D, Chen H, Yan W, Yang D, Chen G, et al. Clinical characteristics of 113 deceased patients with coronavirus disease 2019: retrospective study. BMJ. 2020;368:m1091.

10. Bennett TD, Hayward KN, Farris RW, Ringold S, Wallace CA, Brogan TV. Very high serum ferritin levels are associated with increased mortality and critical care in pediatric patients. Pediatr Crit Care Med. 2011;12(6):e233–6.

11. Liu Z, Sun R, Li J, Cheng W, Li L. Relations of Anemia With the All-Cause Mortality and Cardiovascular Mortality in General Population: A Meta-Analysis. Am J Med Sci. 2019;358(3):191–9.

12. Kernan KF, Carcillo JA. Hyperferritinemia and inflammation. Int Immunol. 2017;29(9):401–9.

13. Wessling-Resnick M. Crossing the Iron Gate: Why and How Transferrin Receptors Mediate Viral Entry. Annu Rev Nutr. 2018;38:38–431.

14. Cassat James E, Skaar Eric P. Iron in Infection and Immunity. Cell Host & Microbe. 2013;13(5):509–19.

15. Wessling-Resnick M. Crossing the Iron Gate: Why and How Transferrin Receptors Mediate Viral Entry. Annual Review of Nutrition. 2018;38(1):431–58.

16. Ganz T. Anemia of Inflammation. New England Journal of Medicine. 2019;381(12):1148–57.

17. Muka T, Glisic M, Milic J, Verhoog S, Bohlius J, Bramer W, et al. A 24-step guide on how to design, conduct, and successfully publish a systematic review and meta-analysis in medical research. Eur J Epidemiol. 2020;35(1):49–60.

18. Huang Y, Tu M, Wang S, Chen S, Zhou W, Chen D, et al. Clinical characteristics of laboratory confirmed positive cases of SARS-CoV-2 infection in Wuhan, China: A retrospective single center analysis. Travel Med Infect Dis. 2020:101606.

19. Xu T, Huang R, Zhu L, Wang J, Cheng J, Zhang B, et al. Epidemiological and clinical features of asymptomatic patients with SARS-CoV-2 infection. J Med Virol. >2020.

20. Liu Y, Du X, Chen J, Jin Y, Peng L, Wang HHX, et al. Neutrophil-to-lymphocyte ratio as an independent risk factor for mortality in hospitalized patients with COVID-19. J Infect. 2020.

21. Li X, Xu S, Yu M, Wang K, Tao Y, Zhou Y, et al. Risk factors for severity and mortality in adult COVID-19 inpatients in Wuhan. J Allergy Clin Immunol. 2020.

22. Lomholt FK, Laulund AS, Bjarnason NH, Jorgensen HL, Godtfredsen NS. Meta-analysis of routine blood tests as predictors of mortality in COPD. Eur Clin Respir J. 2014;1.

23. Rasmussen L, Christensen S, Lenler-Petersen P, Johnsen SP. Anemia and 90-day mortality in COPD patients requiring invasive mechanical ventilation. Clin Epidemiol. 2010;3:3–1.

24. Groenveld HF, Januzzi JL, Damman K, van Wijngaarden J, Hillege HL, van Veldhuisen DJ, et al. Anemia and mortality in heart failure patients a systematic review and meta-analysis. J Am Coll Cardiol. 2008;52(10):818–27.

25. Li Z, Zhou T, Li Y, Chen P, Chen L. Anemia increases the mortality risk in patients with stroke: A meta-analysis of cohort studies. Sci Rep. 2016;6:26636.

26. Shoenfeld Y. Corona (COVID-19) time musings: Our involvement in COVID-19 pathogenesis, diagnosis, treatment and vaccine planning. Autoimmun Rev. 2020:102538.

27. Drakesmith H, Prentice A. Viral infection and iron metabolism. Nat Rev Microbiol. 2008;6(7):541–52.

28. Parrow NL, Fleming RE, Minnick MF. Sequestration and scavenging of iron in infection. Infect Immun. 2013;81(10):3503–14.

